# Transcriptomic Reproducibility in Endometrial Biopsies: Evidence Across Cycles and Tissue Regions in Hormonal Replacement Therapy Endometrial Preparation Protocols

**DOI:** 10.64898/2026.09.23.26363653

**Authors:** Asunta Martinez-Martinez, Patricia Sebastian-Leon, Antonio Parraga-Leo, Pilar Alama, Laura Caracena, Francisco Jose Sanz, Patricia Diaz-Gimeno

## Abstract

**Study question:** Are endometrial gene expression profiles reproducible across hormone replacement therapy (HRT) cycles and across different regions of the same endometrial biopsy?

**Summary answer:** Endometrial transcriptomic profiles were highly reproducible across HRT cycles and across non-adjacent and adjacent regions of the same biopsy.

**What is known already:** Endometrial transcriptomic analyses are increasingly used to understand endometrial physiology and to evaluate endometrial reproductive potential. However, this approach requires an endometrial biopsy in a prior endometrial cycle to the embryo transfer cycle in the IVF treatment. Therefore, endometrial transcriptomic analyses rely on the assumption that the gene expression profile is representative of subsequent endometrial cycles and of the whole biopsy specimen. However, inter-cycle and intra-biopsy reproducibility remain insufficiently characterized.

**Study design, size, duration:** This was a prospective, single-center, observational clinical study including 20 participants recruited between June 2024 and March 2025.

**Participants/materials, setting, methods:** Participants underwent two different endometrial preparation cycles under the same HRT conditions at a private fertility clinic, and an endometrial biopsy was collected during both cycles, with the same duration of progesterone exposure, in the mid-secretory phase by the same clinician. RNA was processed for whole endometrial transcriptome analysis using RNA sequencing. Three paired comparisons were performed: biopsies from the same participant collected in different cycles, non-adjacent regions of the same biopsy, and adjacent regions of the same biopsy. Gene expression reproducibility was assessed using Pearson correlation, hierarchical clustering, principal component analysis, and paired differential gene expression analysis. The reproducibility of estimated cell-type proportions was also assessed by deconvolution to observe potential differences in cellular composition.

**Main results and the role of chance:** In the inter-cycle endometrial gene expression comparison, the mean gene expression correlation coefficient within participant pairs was 0.974 ± 0.012, significantly higher than that observed between participant pairs (*p* = 1.56 × 10^−6^), the mean Euclidean distance was significantly lower within than between participant pairs (*p* = 2.24 × 10^−6^), and no differentially expressed genes were identified within participant pairs. Principal component analysis and hierarchical clustering further supported the similarity of most paired samples. Intra-biopsy comparisons showed even higher reproducibility, with both non-adjacent and adjacent biopsy regions showing mean correlation coefficients of 0.986 ± 0.003 and 0.986 ± 0.002, respectively. The correlation coefficients were significantly higher than those observed between biopsy pairs (*p* = 9.91 × 10^−12^ and *p* = 2.14 × 10^−12^, respectively). No differentially expressed genes were detected within intra-biopsy pairs, all paired regions clustered together by hierarchical clustering and principal component analysis, and intra-biopsy pairs showed lower Euclidean distances than inter-biopsy pairs (*p =* 9.91 × 10^−12^ and *p* = 2.14 × 10^−12^ in non-adjacent and adjacent regions, respectively). Estimated cell-type proportions did not differ significantly between paired samples in any of the three comparisons.

**Limitations, reasons for caution:** The findings are specific to samples analyzed using the AmpliSeq Transcriptome Human Gene Expression Panel protocol. Although other transcriptomic platforms should be tested, we are expecting the same reproducibility due to the consistency of RNA seq platforms. All biopsies were collected by the same clinician, so operator-related variability could not be directly assessed. These findings can be extrapolated only to HRT cycles.

**Wider implications of the findings:** These findings support the reproducibility of endometrial gene expression profiles under controlled HRT conditions from biopsies collected by the same clinician and suggest that a single biopsy region can provide a representative molecular profile of the whole biopsy specimen. These findings strengthen the methodological basis for bulk endometrial RNA sequencing studies and support the reliability of transcriptomic-based tools for endometrial evaluation in an endometrial cycle prior to an embryo transfer cycle for reproductive medicine.

**Funding:** This study was supported by the IVI Foundation (2201-FIVI-004-PD). Patricia Diaz-Gimeno was supported by the Instituto de Salud Carlos III (ISCIII) through the Miguel Servet program (CP20/00118) co-funded by the European Union. Asunta Martinez-Martinez and Francisco Jose Sanz are funded by the Instituto de Salud Carlos III (ISCIII) through a predoctoral fellowship program (FI24/00104 [A.M.-M.]) and the Sara Borrell postdoctoral program (CD23/00032 [F.J.S.]) co-funded by the European Union.

**Disclosures:** None to declare.

**Trial registration number:** Not applicable.

**LAY SUMMARY:** *What does this mean for patients?:* The endometrium is the inner lining of the womb where embryo implantation occurs, and it supports early pregnancy. In fertility research, small samples (biopsies) of endometrial tissue are often tested to study endometrial health and to develop tools for assessing reproductive chances. During in vitro fertilization treatment, these tests are usually performed on a biopsy collected in a prior menstrual cycle than the cycle when embryo transfer occurs. The tests also usually analyze only a small part of the biopsy. Therefore, it is important to know whether the results from one cycle are similar to the next cycles, and whether one small part of the biopsy represents the whole sample. In this study, we analyzed endometrial biopsies collected from volunteers from two hormone-prepared cycles. We also compared different regions of the same biopsy. We found that the expressed genes were highly similar between cycles from the same woman and between different regions of the same biopsy. The estimated proportions of the main cell types in the tissue were also stable. These results suggest that, under the controlled conditions used in this study, endometrial biopsies provide a consistent picture of the endometrium across menstrual cycles. This finding supports the reliability of endometrial gene expression-based studies and tools.

## INTRODUCTION

The endometrium, the innermost layer of the uterus, is a hormonally regulated and highly dynamic tissue that undergoes cyclic remodeling throughout the menstrual cycle. These changes are essential for establishing a functional environment for embryo implantation and supporting early pregnancy (Critchley *et al*., 2020). Given that the endometrium has a central role in fertility, numerous studies have focused on endometrial function by using transcriptomic analyses of endometrial biopsies for modelling complexity and biomarker discovery (Altmäe *et al*., 2017; Bastu *et al*., 2019; Devesa-Peiro *et al*., 2022; Diaz-Gimeno *et al*., 2022, 2024; Díaz-Gimeno *et al*., 2011; Koot *et al*., 2016; Shi *et al*., 2018). In addition, transcriptomic analyses of endometrial biopsies have been translated into the clinical setting to evaluate endometrial function optimizing embryo transfer timing and tools intended to estimate endometrial reproductive prognosis (Altmäe *et al*., 2017; Diaz-Gimeno *et al*., 2024; Díaz-Gimeno *et al*., 2011, 2017; Enciso *et al*., 2018; Hamamah and Delphine, 2016). However, endometrial biopsy is an invasive procedure that does not allow for embryo transfer in the same cycle, requiring a previous independent evaluation cycle. Consequently, clinicians rely on the assumption that endometrial physiology remains stable across consecutive cycles, inferring the molecular state of the next cycle for embryo transfer

Transcriptomics reproducibility across menstrual cycles has been analyzed in several works. Díaz-Gimeno and colleagues evaluated the accuracy and reproducibility of the endometrial receptivity array (ERA) gene expression platform, which is based on the expression of a 238-gene signature, by comparing ERA diagnoses obtained from two endometrial biopsies collected from the same women in different cycles (Díaz-Gimeno *et al*., 2013). The authors reported concordant ERA diagnoses across all biopsy pairs, and samples from the same individuals clustered together in principal component analysis (PCA). However, only seven biopsy pairs were analyzed, and although the pairs in the same patient corresponded to the same phase of the menstrual cycle, not all the patients participated in the same menstrual phase. Therefore, it is not possible to elucidate if samples from the same patient clustered together due to they belonged to the same patient or because the differences in the menstrual phase between patients. Similarly, Evans and colleagues investigated inter-cycle variability in the expression of 38 genes using microarray analysis of laser-microdissected endometrial glands and stroma from six women (Evans *et al*., 2018). They reported that approximately 60% of the analyzed genes exhibited less than 30% variation between cycles. Although these previous studies pointed out inter-cycle reproducibility, both studies were limited by small sample sizes, which may reduce statistical power, and by the restricted number of genes analyzed, precluding a comprehensive assessment of transcriptomic variability across the entire endometrial transcriptome. Moreover, these studies relied on microarray technology, which has largely been superseded by RNA sequencing (RNA-seq) in transcriptomic research over the past decade.

Because of the importance of reporting transcriptomic reproducibility cycle by cycle, which will provide translational value, a 2026 study evaluated the whole endometrial transcriptome, rather than a reduced set of genes, and reported that intra-individual transcriptomic variance was significantly lower than inter-individual transcriptomic variance (Teh et al., 2026). However, this study was conducted using endometrial biopsies obtained in natural cycles from women aged 26 to 45 years (36.8 ± 4.6 years), which introduces more natural interpersonal variability in endometrial progression than hormone replacement therapy (HRT) cycles do, because hormone levels in these HRT cycles are controlled (Alfer *et al*., 2022). Therefore, biopsies obtained in two natural cycles might be more affected by this variability in hormone levels. This consideration is particularly relevant for patients of advanced maternal age undergoing in vitro fertilization (IVF), in whom HRT cycles are commonly used for endometrial preparation, especially for oocyte receptors treatments (Coughlan *et al*., 2023; de Ziegler *et al*., 1991) and whose representation in reproductive medicine is progressively increasing (Seshadri *et al*., 2021). This evidence reinforces the need for assessing endometrial transcriptomic reproducibility under HRT conditions.

It is well established that the relative abundance of the different cell populations found in endometrial tissue varies across the phases of the menstrual cycle (Garcia-Alonso *et al*., 2021), and differences in cell-type proportions substantially alter the biological interpretations derived from differential expression analyses of bulk endometrial transcriptomic data (Suhorutshenko *et al*., 2018). In this context, Teh and colleagues reported that cell-type proportions remained stable between biopsies collected at the same endometrial phase in different natural cycles (Teh *et al*., 2026). However, it remains unclear whether endometrial cellular composition is conserved across the same endometrial phase in different HRT cycles within the same woman. Evidence regarding the consistency of cell-type proportions across menstrual cycles is limited.

Additionally, transcriptomic analyses are typically performed on a small, non-specified region of the biopsy, implicitly assuming spatial homogeneity of the endometrium and negligible variability in gene expression across different regions of the tissue. To date, neither transcriptomic reproducibility nor cell-type proportion consistency across different regions within the same biopsy has been demonstrated. This gap may directly affect the interpretation and extrapolation of findings from bulk endometrial transcriptomic studies.

Also, it has to be considered the advancement in transcriptomics techniques by RNA sequencing in the last decade, a technique that is sensitive to RNA quality and degradation in clinical biopsy-derived samples, which enables robust whole-transcriptome analysis in contrast to targeted approaches or microarray-based analyses (Fasold and Binder, 2014). Moreover, HRT cycles are widely used in women undergoing IVF, such as in patients of advanced maternal age with premenopausal symptoms; thus, evaluating HRT cycle reproducibility is necessary, as it has not yet been specifically assessed. Therefore, the aim of this study was to evaluate the entire endometrial gene expression and cell-type proportion consistency by RNA-seq across HRT cycles and different endometrial biopsy regions.

## MATERIALS AND METHODS

### Ethical approval

This study was approved by the ethics committee of the Instituto Valenciano de Infertilidad (Valencia, Spain; 2201-FIVI-004-PD) and adhered to the fundamental principles set forth in the Declaration of Helsinki, the Council of Europe Convention on Human Rights and Biomedicine, and the UNESCO Universal Declaration on the Human Genome and Human Rights. It also complied with the requirements established in Spanish legislation in the fields of biomedical research, personal data protection, and bioethics. Written informed consent was obtained from all recruited participants.

### Study design

The study design is presented in Figure 1.

**Figure 1.**
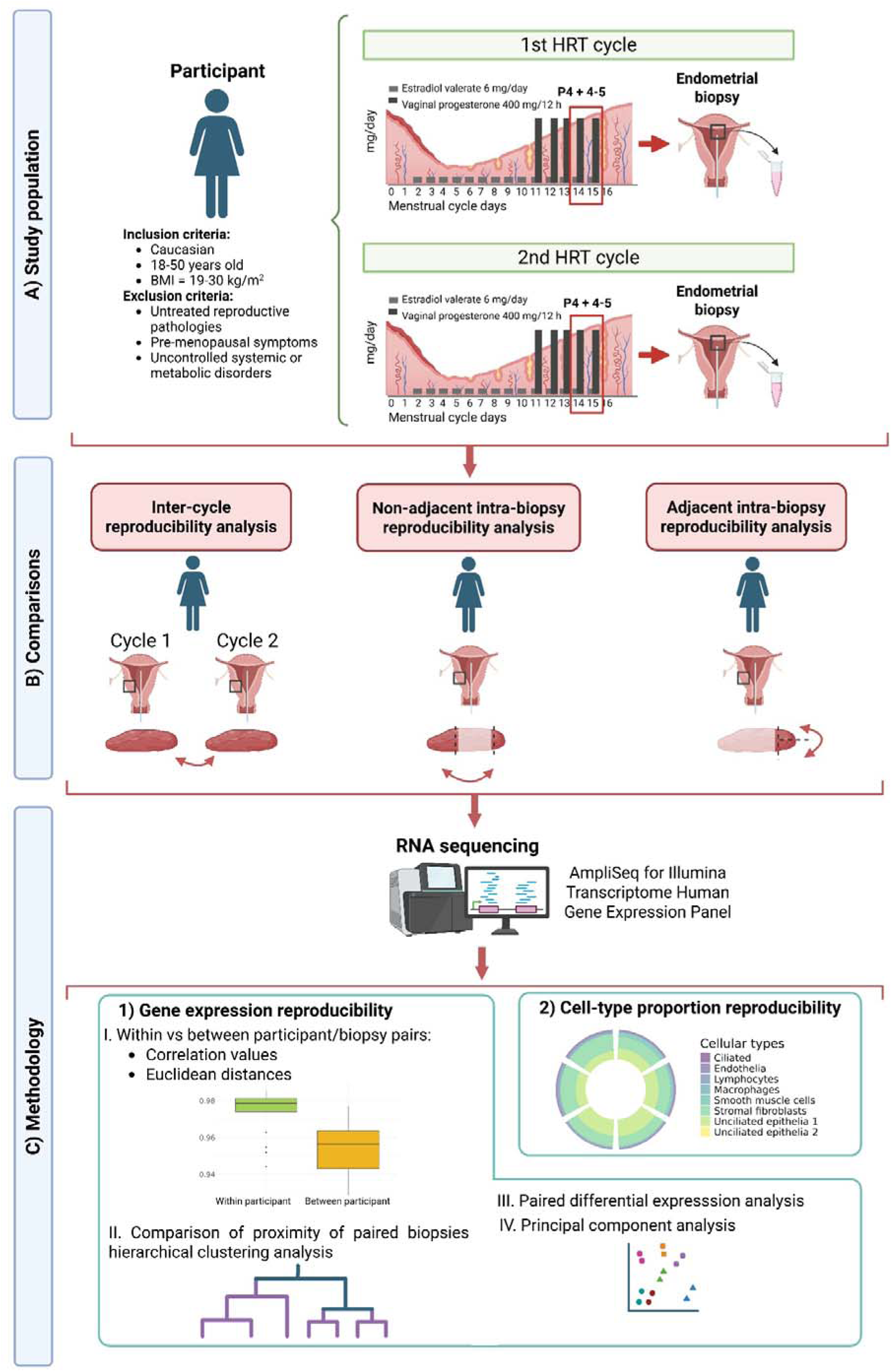
Study design. **A) Study population.** This study included women aged 18–50 years, with a BMI between 19 and 30 kg/m², who were Caucasian, and had no untreated reproductive pathologies, severe pre-menopausal symptoms, or uncontrolled systemic or metabolic disorders. These participants underwent two hormone replacement therapy (HRT) cycles. HRT was initiated on day 2–3 of the menstrual cycle with oral estradiol valerate at a dose of 6 mg/day. Once an endometrial thickness of at least 6.5 mm was achieved and serum progesterone levels were below 1 ng/mL, vaginal progesterone was initiated at a dose of 400 mg every 12 hours and maintained until the day of biopsy. Endometrial biopsies were collected on the 4^th^ or 5^th^ day of progesterone exposure (P4+5). **B) Comparisons.** Using all collected samples, three comparative analyses were performed based on the endometrial transcriptome of each biopsy. First, endometrial biopsies obtained from the same women in two HRT cycles were compared (inter-cycle comparison). Second, two non-adjacent regions from the same biopsy were compared (non-adjacent intra-biopsy comparison). Third, two adjacent regions from the same biopsy were compared (adjacent intra-biopsy comparison). **C) Methodology.** Samples RNA was sequenced using AmpliSeq for Illumina Transcriptome Human Gene Expression Panel protocol. **1.** For each comparison, gene expression reproducibility was evaluated: I) Correlation values and Euclidean distances were compared within and between participant/biopsy pairs, II) proximity of paired biopsies was compared by hierarchical clustering, III) paired differential expression analysis was performed, and IV) principal component analysis was performed. **2**. Changes in cell-type proportions were assessed by performing cell-type deconvolution for each sample, followed by pairwise comparison. The cell types analyzed were ciliated epithelial cells, endothelia cells, lymphocytes, macrophages, smooth muscle cells, stromal fibroblasts and unciliated epithelia cells types 1 and 2.

This study included two endometrial biopsies from 20 participants collected during two mid-secretory endometrial cycles under the same conditions of HRT (Figure 1A). The HRT cycle started on day 2 or 3 of menstruation, when participants received oral estradiol valerate at a dose of 6 mg/day until the endometrial biopsy. Approximately 9–14 days after the start of estrogen therapy, if the endometrial thickness was at least 6.5 mm and showed a trilaminar pattern, the same estradiol dose was maintained. If the endometrium was considered insufficiently developed, the estradiol dose was increased, and the participant was reassessed 5–7 days later. Once an endometrial thickness of at least 6.5 mm was achieved and serum progesterone levels were below 1 ng/mL, vaginal progesterone was initiated at a dose of 400 mg every 12 hours and maintained until the day of biopsy. If serum progesterone levels exceeded 1 ng/mL before progesterone administration, the cycle was cancelled. The endometrial biopsy was collected during the mid-secretory phase.

Biopsies were analyzed to assess gene expression reproducibility and cell-type proportion variability in the human endometrium across HRT cycles and across different regions within the same biopsy. The transcriptomes of the endometrial biopsies were sequenced, and three pairwise comparisons were performed. First, endometrial biopsies obtained from the same women in two HRT cycles under the same conditions were compared to evaluate inter-cycle variability. Second, two non-adjacent regions from the same biopsy were compared to assess spatial variability within a single biopsy. Third, two adjacent regions from the same biopsy were compared to evaluate local tissue reproducibility (Figure 1B). For each comparison, gene expression reproducibility was evaluated by comparing correlation values and Euclidean distances within and between participant/biopsy pairs, comparing proximity of paired biopsies by analyzing hierarchical clustering, and performing paired differential expression analysis and PCA (Figure 1C). In addition, changes in cell-type proportions were assessed by performing cell-type deconvolution for each sample, followed by pairwise comparison.

### Participants and clinical follow-up

Participants (n = 20) were recruited for a prospective, single-center, observational study between June 2024 and March 2025 at a private fertility clinic in Valencia, Spain. Participants met the following inclusion criteria: Caucasians, 18–50 years old, body mass index of 19–30 kg/m^2^, and with an endometrial thickness of more than 6.5 mm with trilaminar structure in the proliferative phase. Exclusion criteria were untreated reproductive pathologies that may compromise endometrial function, severe premenopausal symptoms, and uncontrolled systemic or metabolic disorders. Baseline participant characteristics, including age and body mass index, the duration of progesterone exposure at the time of biopsy collection, and the number of menstrual cycles between biopsies, were obtained from internal medical records.

### Endometrial biopsy collection, RNA sequencing, and data processing

All participants underwent two successive HRT cycles, and all endometrial biopsies were obtained in the mid-secretory phase in each cycle. Endometrial biopsies were obtained from the uterine fundus using a cannula (Pipelle CCD, CCD Laboratories, Paris, France) under sterile conditions. The biopsies were obtained by the same clinician using the same procedure. Anonymized samples were stored in RNAlater (Sigma-Aldrich, Madrid, Spain) at−80°C. RNA was extracted using the miRNeasy Mini Kit (Qiagen, Hilden, Germany) following the manufacturer’s instructions. A commercial control sample (Universal Human Reference RNA, Agilent Technologies, Santa Clara, CA, USA), composed of RNA from multiple human tissues and cell lines, was included in duplicate as an external reference to contextualize the magnitude of the observed correlation coefficients. RNA quality was assessed using the NanoDrop One (AF-00342, Thermo Fisher Scientific, Waltham, MA, USA) and the 4200 TapeStation System (Agilent Technologies). Only samples that met the following RNA quality criteria were included in the study: 260/280 ratio, ∼2.0; 260/230 ratio,

1.8–2.2; RNA integrity number, ≥3; and RNA fragments with more than 200 nucleotides, ≥70%. Samples meeting quality criteria were sequenced using the AmpliSeq for Illumina Transcriptome Human Gene Expression Panel protocol (Illumina, San Diego, CA, USA) on a NextSeq 500/550 system (Illumina) using a paired-end design with 150 cycles and sequencing 10 million reads per sample. Raw data were evaluated using FastQC (version 0.11.9) (Andrews *et al*., 2012). STAR (version 2.7.3) (Dobin *et al*., 2013) was used to align high-quality data using GRCh37/hg19 as a reference (Genome Reference Consortium, 2009). Gene counts were obtained using featureCounts (version 2.0.3) (Liao *et al*., 2014), and low-quality counts (Q < 30) were filtered. Genes with less than 1.5 counts per million in more than 95% of the samples were filtered out; the counts per million calculation was performed using the edgeR R package (version 3.32.1) (Chen *et al*., 2016). Thereafter, counts were normalized using voom transformation and quantile normalization using the limma R package (version 3.46.0) (Ritchie *et al*., 2015). Outliers were detected based on Hotelling’s distance using the HotellingEllipse R package (version 1.2.0) (Brereton, 2016) with a 95% confidence level. Possible batch effects were detected using principal variance component analysis (pvca R package, version 1.50.0) (Bushel, 2025), and variables explaining more than 10% of the total variance were considered potential batch factors. Identified batch effects were subsequently corrected using linear models implemented in the limma R package.

### Gene expression reproducibility across cycles and biopsy regions

Gene expression reproducibility was evaluated across all comparisons (inter-cycle, and non-adjacent and adjacent intra-biopsy) following the same strategy. First, a Pearson correlation analysis using the stats R package (version 4.5.2) (R Core Team, 2021) was performed to measure the degree of similarity between gene expression profiles across all samples of each comparison. Correlation values obtained within and between participant/biopsy pairs were compared using a non-parametric Wilcoxon test. Second, paired differential expression analysis was performed to identify genes differentially expressed within and between participant/biopsy pairs using the limma R package. Finally, PCA and hierarchical clustering based on Euclidean distance were conducted for all samples within each comparison to determine whether within participant/biopsy pairs clustered together, reflecting similar transcriptomic profiles. PCA and hierarchical clustering were performed using the stats R package. We compared the Euclidean distance within and between participant/biopsy pairs using a Wilcoxon test from the stats R package.

### Cell-type proportion reproducibility across cycles and biopsy regions

Cell-type proportions were estimated using reference-based bulk RNA-seq deconvolution. A single-cell transcriptomic dataset of 19 endometrial biopsies from GSE111976 was retrieved from the Gene Expression Omnibus (Wang *et al*., 2020) and analyzed using Seurat (version 5.3.0) (Hao *et al*., 2024). Only dataset samples collected in the mid-secretory phase were included so as to match the endometrial phase criteria of the present study. After quality control, a cell type-specific signature matrix was generated and used as a reference to estimate cell-type proportions from the study samples using the DWLS R package (version 0.1.0) (Tsoucas *et al*., 2019). Linear mixed-effects models were used to evaluate differences in cell-type proportions within and between participant/biopsy pairs, using paired participant as a random effect.

### Statistical analysis

Participants’ baseline clinical characteristics are described as mean ± standard deviation (SD). All statistical analyses were conducted in R. Graphical results were generated with the ggplot2 R package (version 3.5.1) (Wickham, 2016). In all comparisons, *p* values or false discover rates of less than 0.05 were considered statistically significant.

## RESULTS

### Clinical characterization of participants

The participants are 36.0 ± 6.5 years, with a body mass index of 23.4 ± 3.1 kg/m^2^, and a duration of progesterone exposure of 111.9 ± 7.6 hours at the time of biopsy collection. Of the 20 recruited participants, 17 women were included for the inter-cycle and adjacent intra-biopsy reproducibility analyses, and 16 women were included for the non-adjacent intra-biopsy reproducibility analysis. Samples not included in the final analyses were excluded due to a volunteer drop-out, insufficient tissue to perform all comparisons, outlier removal, or a difference in duration of progesterone exposure greater than 24 hours within participant pairs, which was used to avoid transcriptomic differences driven by endometrial timing rather than by true inter-cycle variability. More details are in Supplementary Figure S1.

The number of cycles between biopsies collected from the same woman ranged from 1 to 6 menstrual cycles with a mean of 2.6 cycles, and the difference in duration of progesterone exposure within participant biopsies ranged from 0.9 to 23.9 hours. Potential batch effects in gene expression related to experimental variables were not detected (Supplementary Figure S2).

### High transcriptomic reproducibility across cycles and biopsy regions

In the inter-cycle reproducibility analysis, the mean gene expression correlation coefficient within participant pairs was 0.974 ± 0.012. This value is similar to the correlation coefficient obtained between positive control replicates (0.984), indicating strong transcriptomic reproducibility across cycles in the same woman, and is significantly higher than the mean correlation coefficient observed between participant pairs (*p* = 1.57 × 10^-6^) (Figure 2A). Consistent with these findings, no gene expression differences were identified within participant pairs (adj *p* > 0.355). The mean Euclidean distance within participant pairs was significantly lower than between participant pairs (*p* = 2.24 × 10^-6^) (Figure 2A) showing inter-cycle reproducibility. In addition, techniques aimed at assessing patterns of transcriptomic similarity and heterogeneity revealed that paired samples clustered together, supporting their similar transcriptomic behavior. However, the two samples of the same pair not clustering together might be driven by factors not related with the inter-cycle variability since they do not cluster with the rest of the cohort. To see more details about how samples are more similar within than between participants is detailed in Supplementary Figure S3A.

**Figure 2.**
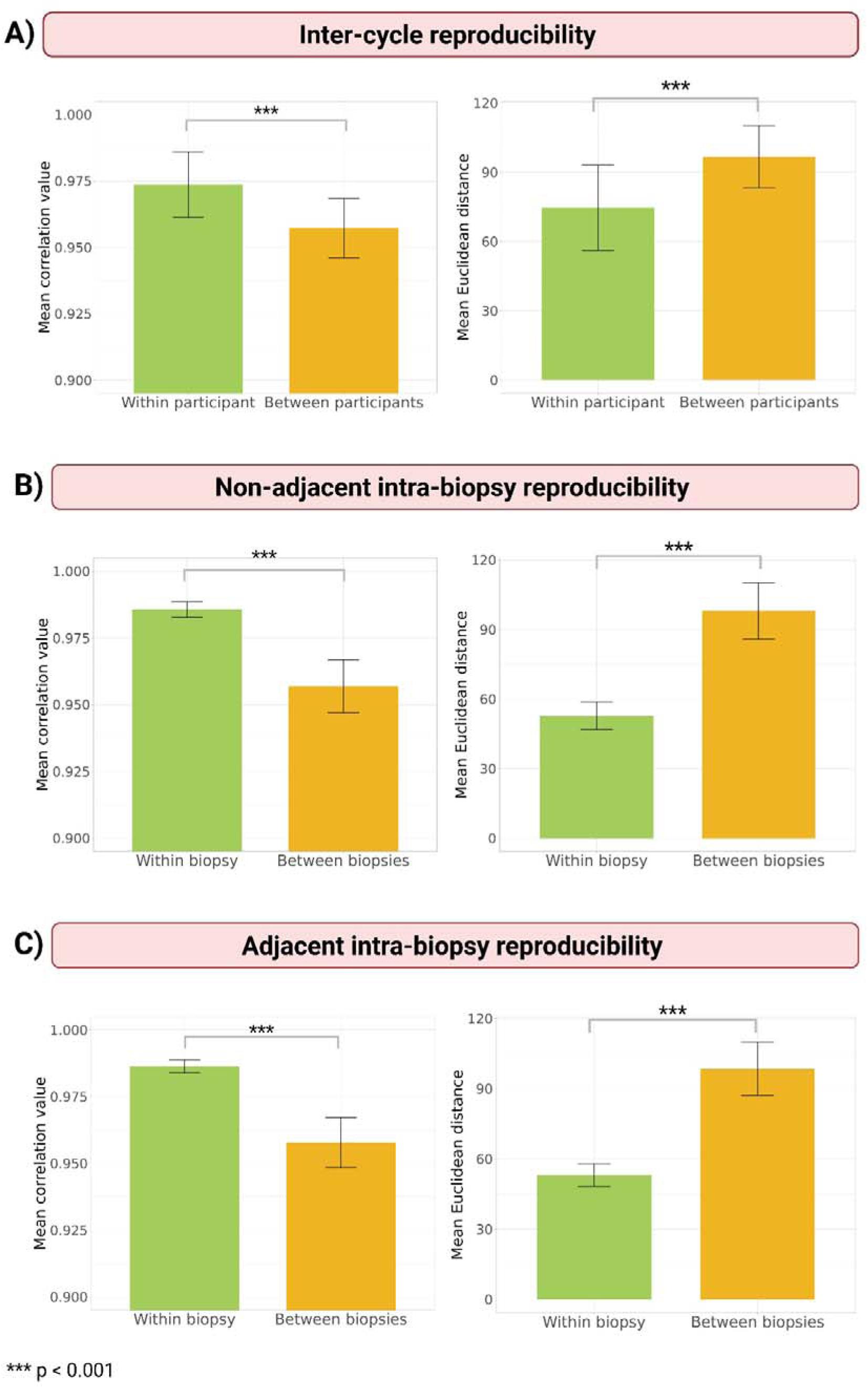
Gene expression reproducibility for endometrial transcriptomic analysis. Bar plots represent correlation coefficients and Euclidean distances for (A) inter-cycle, (B) non-adjacent intra-biopsy, and (C) adjacent intra-biopsy comparisons, within-participant/biopsy pairs were compared with between-participant/biopsy pairs. In all comparisons, mean correlation coefficients were higher in within-participant or within-biopsy pairs than in between-participant or between-biopsy pairs, whereas mean Euclidean distances were lower. In both intra-biopsy comparisons, the standard deviation (SD) of within-biopsy pairs was lower than that of between-biopsy pairs for both correlation coefficients and Euclidean distances. In the inter-cycle comparison, SD values were similar between groups. Data are presented as mean ± SD. Significant differences were observed in all comparisons (***p < 0.001).

As expected, the intra-biopsy comparisons demonstrated even higher concordance. Both non-adjacent and adjacent biopsy regions showed high correlation with their corresponding paired region (mean correlation coefficients of 0.986 ± 0.003 and 0.986 ± 0.002 in non-adjacent and adjacent biopsy regions, respectively), reflecting near-perfect transcriptomic reproducibility within the same biopsy specimen. These values are significantly higher than the mean correlation coefficient observed between biopsy pairs (*p* = 9.91 × 10^-12^ and *p* = 2.14 × 10^-12^ in non-adjacent and adjacent biopsy regions, respectively) and also had lower standard deviations (Figure 2B, 2C). Consistently, no significantly differentially expressed genes were detected within biopsy pairs in either intra-biopsy analysis (adj *p* > 0.081). Additionally, the mean Euclidean distance was significantly lower within biopsy pairs than between biopsy pairs (*p* = 9.91 × 10^-12^ and *p* = 2.14 × 10^-12^ in non-adjacent and adjacent biopsy regions, respectively) and also had lower standard deviations (Figure 2B, 2C). The higher gene expression similarities within participants were consistent with results using other multiparametric approaches, with all intra-biopsy regions clustering closer together than for inter-cycle gene expression comparisons (Supplementary Figure S3A-C).

### No differences in cell-type proportions among cycles and biopsy regions

To assess whether there are different cell-type proportions between pairs, we performed deconvolution analysis on the RNA-seq data. The estimated proportions of each cell type included in the analysis were compared within biopsy pairs and between participant/biopsy pairs across the three comparisons (Figure 3), with no statistically significant differences identified (all false discovery rates were greater than 0.19) (Supplementary Table S1). These results indicate that the endometrial cellular composition remains similar between cycles from the same women and between non-adjacent and adjacent biopsy regions, supporting the biological stability and reproducibility of the endometrial biopsies.

**Figure 3.**
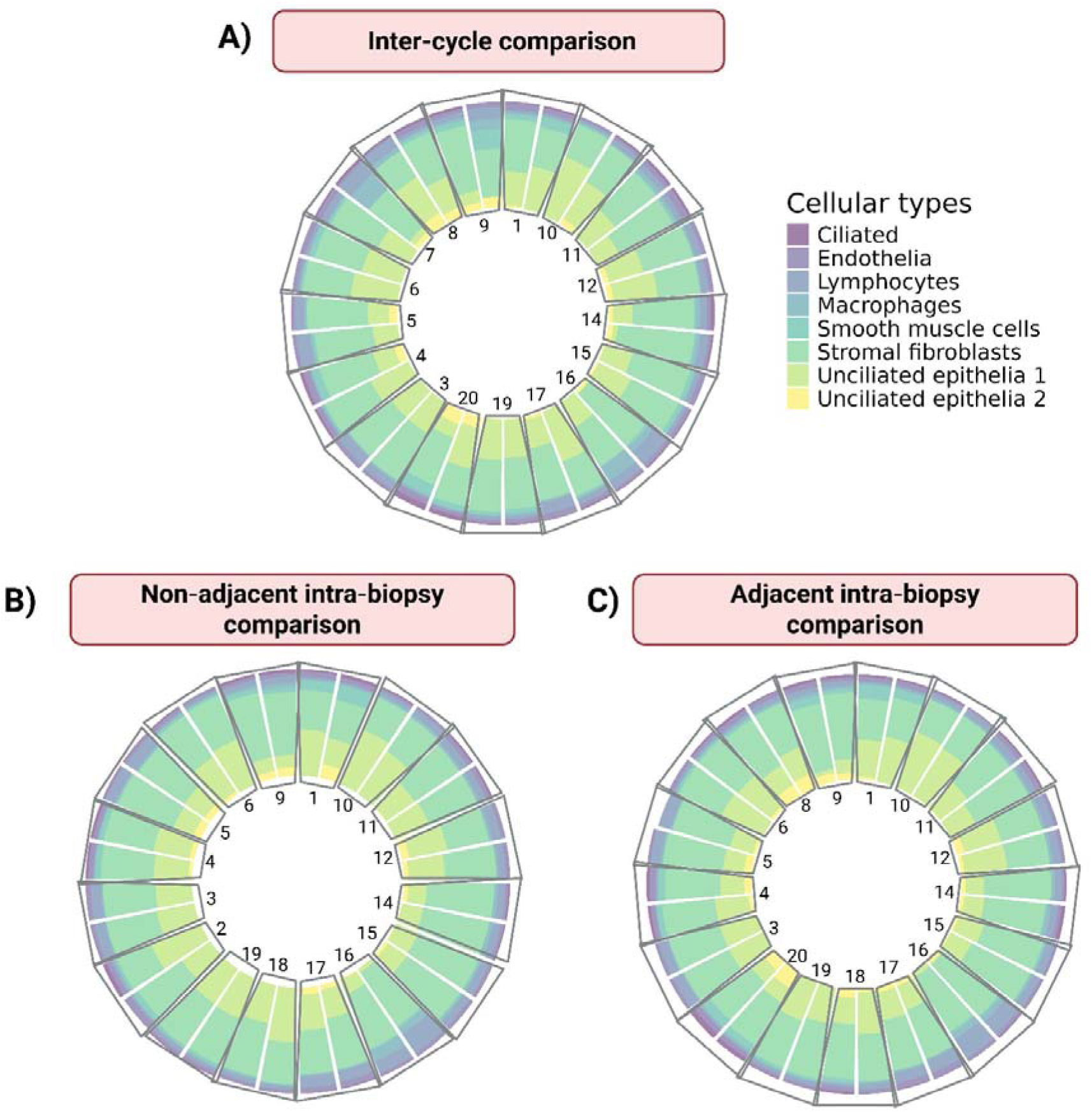
Cell-type proportions between participant pairs and within biopsy pairs. The cell-type proportions were obtained from deconvolution analysis and grouped by pair; each number represents each pair. Each sample pair presented similar cell-type proportions. Statistically insignificant differences (p > 0.05, false discovery rate > 0.05) were observed for each cell-type proportion for paired samples for all comparisons.

## DISCUSSION

Here, we measured the reproducibility of endometrial transcriptomic profiles and cell-type proportions across cycles under the same HRT conditions. Our findings demonstrated that the gene expression remains stable in biopsies collected by the same clinician from the same women (inter-cycle comparison) and across distinct regions within the same endometrial biopsy (non-adjacent and adjacent intra-biopsy comparison). For the inter-cycle comparison, the biopsies had strong transcriptomic similarity as highlighted by several approaches, with paired samples clustering together and displaying higher gene expression correlation, shorter Euclidean distance, and no differentially expressed genes. Intra-biopsy comparisons had even higher concordance, as expected, as both non-adjacent and adjacent biopsy regions had near-perfect transcriptomic correlation and lower Euclidean distance compared with random pairs and clustered consistently with their corresponding paired regions. No differentially expressed genes were identified between paired biopsy regions, supporting the presence of limited spatial transcriptomic variability within the same biopsy specimen. In addition, deconvolution analysis revealed no significant differences in the estimated proportions of the analyzed cell types, either between HRT cycles or between different regions of the same biopsy. Together, these findings suggest that, under standardized HRT conditions and with biopsies collected by the same clinician, both the bulk transcriptomic profile and the inferred cellular composition of mid-secretory endometrial biopsies remain highly stable across cycles and across biopsy regions.

Only a limited number of studies have investigated inter-cycle reproducibility in the endometrium, and their findings are constrained by several limitations, including small sample sizes, sample collection only during natural menstrual cycles, and the evaluation of a limited number of RNA-seq platforms (Díaz-Gimeno *et al*., 2013; Evans *et al*., 2018; Teh *et al*., 2026). Results from biopsies from natural cycles could be more variable, especially in women older than 40 years for whom premenopausal symptoms are associated with gradual hormone level changes; in those cases, biopsy timing is less tightly controlled than in HRT cycles, potentially leading to greater variability among cycles. Our study addressed these limitations by assessing whole-transcriptome reproducibility in endometrial biopsies collected under controlled HRT conditions, using a standardized RNA-seq approach that had not been previously evaluated for this purpose, and including a sample size determined by statistical sample size estimation, which was also relatively larger than those of previous studies (Díaz-Gimeno *et al*., 2013; Evans *et al*., 2018).

Furthermore, our findings are consistent with those of previous studies that report associations between endometrial gene expression profiles and reproductive outcomes in the embryo transfer cycle after biopsy collection (Diaz-Gimeno *et al*., 2024; Díaz-Gimeno *et al*., 2017; Koot *et al*., 2016; Parraga-Leo *et al*., 2023; Sanchez-Reyes *et al*., 2025). If endometrial transcriptomic profiles were not reproducible across cycles, the molecular information obtained from a biopsy would be less likely to reflect reproductive outcomes in a subsequent cycle. Consistently, results reported by these works supported the clinical relevance of these transcriptomic signatures, reinforcing the inter-cycle reproducibility observed in the present study.

Additionally, our study is the first one to evaluate reproducibility across different regions within the same endometrial biopsy, making these findings particularly valuable for the interpretation and translation of bulk endometrial transcriptomic analyses in HRT cycles. The high intra-biopsy reproducibility observed in both gene expression and cell-type proportion supports the assumption that tissue heterogeneity does not affect the biopsy result and that a single biopsy region can provide a representative molecular profile of the whole specimen, reducing concerns about sampling-related variability in endometrial transcriptomic studies. Importantly, this finding also contributes to validating the use of transcriptomic-based tools, which commonly rely on the analysis of a limited biopsy region to infer the whole endometrial status.

Estimated cell-type proportions remained stable across different regions of the same biopsy and also across HRT cycles within the same participant, consistent with the findings reported by Teh and colleagues in natural cycles, which only reported this in two different cycles but did not address the comparison between different regions of the same biopsy (Teh *et al*., 2026). These findings are particularly relevant because Suhorutshenko and colleagues showed that differences in the relative proportions of epithelial and stromal cells can lead to different results by differential expression analysis (Suhorutshenko *et al*., 2018). Therefore, the stability of cell-type proportions observed in our study supports the robustness of our transcriptomic comparisons and suggests that the observed reproducibility is not confounded by major differences in cellular proportions.

The AmpliSeq Transcriptome Human Gene Expression Panel protocol was selected for RNA-seq because it allows whole-transcriptome profiling from low-quality RNA samples, a common limitation in endometrial biopsies in the clinical setting, and because it is a standardized, widely used, and cost-effective RNA-seq approach. Nevertheless, as reported by Diaz-Gimeno and colleagues, transcriptomic results may differ depending on the technology used (Diaz-Gimeno *et al*., 2022). Therefore, our findings demonstrate inter-cycle and intra-biopsy reproducibility when samples are analyzed by AmpliSeq, but they should not be directly generalized to all RNA-seq transcriptomic protocols. Further platform-specific studies are needed, although the evidence accumulated so far across the technologies evaluated supports the reproducibility of endometrial transcriptomic profiles by RNA seq (Díaz-Gimeno *et al*., 2013; Evans *et al*., 2018; Teh *et al*., 2026). Another aspect that should be considered is that all biopsies included in this study were collected by the same clinician. Therefore, we cannot exclude the possibility that reproducibility may be affected when biopsies are collected by different operators. However, previous studies that included hundreds of endometrial biopsies collected by different clinicians have shown that this variable does not influence the transcriptomic association of endometrial samples with reproductive outcomes in different cycles (Diaz-Gimeno *et al*., 2022). Thus, although operator-related variability should be acknowledged, the available evidence suggests that endometrial transcriptomic profiles remain robust across biopsy collection settings.

In conclusion, endometrial transcriptomic profiles showed high gene expression reproducibility under the specific clinical and methodological conditions evaluated in this study. Therefore, our findings support that, when these conditions are maintained, endometrial transcriptomic analysis provides a stable and reproducible molecular profile for endometrial evaluation, reinforcing its potential translational value as an endometrial prognostic tool in IVF.

## AUTHORS’ ROLES

P.D.-G. designed the study. A.M.-M., P.A., and L.C. collected the data. A.M.-M., P.S.-L., and A.P.-L. performed the analysis, supervised by P.D.-G. A.M.-M., P.S.-L., A.P.-L., and F.J.S. interpreted the data and drafted the manuscript, supervised by P.D.-G. All authors contributed to the interpretation of the data, revised the manuscript critically for important intellectual content, approved the final version to be submitted and published, and agree to be accountable for all aspects of the work. P.D.-G. acted as an overall guarantor, accepted full responsibility for the work and the study’s conduct, had access to the data, and controlled the decision to publish.

## ACKNOWLEDGMENTS

The authors thank the volunteers who participated; Michelle McRae, PhD, ELS, for editing a presubmission draft of this manuscript; and the Genomic Unit of the Instituto de Investigación Sanitaria La Fe for providing the reagents and infrastructure required for RNA sequencing.

## DATA AVAILABILITY

The data underlying this article will be shared on reasonable request to the corresponding author.

## REFERENCES

Alfer J, Fattahi A, Bleisinger N, Antoniadis S, Krieg J, Dittrich R, Beckmann MW, Hartmann A, Popovici RM, Tremellen K. Individual dynamics of uterine natural killer cells in natural and stimulated cycles monitored using a new endometrial dating method. Am J Reprod Immunol 2022;88:e13620.

Altmäe S, Koel M, Võsa U, Adler P, Suhorutšenko M, Laisk-Podar T, Kukushkina V, Saare M, Velthut-Meikas A, Krjutškov K, et al. Meta-signature of human endometrial receptivity: a meta-analysis and validation study of transcriptomic biomarkers. Sci Rep 2017;7:10077.

Andrews S, Krueger F, Segonds-Pichon A, Biggins L, Krueger C, Wingett S. A quality control tool for high throughput sequence data. 2012. Available at: https://www.bioinformatics.babraham.ac.uk/projects/fastqc/.

Bastu E, Demiral I, Gunel T, Ulgen E, Gumusoglu E, Hosseini MK, Sezerman U, Buyru F, Yeh J. Potential Marker Pathways in the Endometrium That May Cause Recurrent Implantation Failure. Reprod Sci 2019;26:879–890.

Brereton RG. Hotelling’s T squared distribution, its relationship to the F distribution and its use in multivariate space. Journal of Chemometrics 2016;30:18–21.

Bushel P. pvca: Principal Variance Component Analysis (PVCA). 2025. Available at: https://bioconductor.org/packages/release/bioc/html/pvca.html.

Chen Y, Lun ATL, Smyth GK. From reads to genes to pathways: differential expression analysis of RNA-Seq experiments using Rsubread and the edgeR quasi-likelihood pipeline. F1000Res 2016;5:1438.

Coughlan C, Ata B, Gallego RD, Lawrenz B, Melado L, Samir S, Fatemi H. Interindividual variation of progesterone elevation post LH rise: implications for natural cycle frozen embryo transfers in the individualized medicine era. Reprod Biol Endocrinol 2023;21:47.

Critchley HOD, Maybin JA, Armstrong GM, Williams ARW. Physiology of the Endometrium and Regulation of Menstruation. Physiol Rev 2020;100:1149–1179.

Devesa-Peiro A, Sebastian-Leon P, Parraga-Leo A, Pellicer A, Diaz-Gimeno P. Breaking the ageing paradigm in endometrium: endometrial gene expression related to cilia and ageing hallmarks in women over 35 years. Human Reproduction 2022;37:762–776.

Díaz-Gimeno P, Horcajadas JA, Martínez-Conejero JA, Esteban FJ, Alamá P, Pellicer A, Simón C. A genomic diagnostic tool for human endometrial receptivity based on the transcriptomic signature. Fertil Steril 2011;95:50–60, 60.e1–15.

Díaz-Gimeno P, Ruiz-Alonso M, Blesa D, Bosch N, Martínez-Conejero JA, Alamá P, Garrido N, Pellicer A, Simón C. The accuracy and reproducibility of the endometrial receptivity array is superior to histology as a diagnostic method for endometrial receptivity. Fertil Steril 2013;99:508–517.

Díaz-Gimeno P, Ruiz-Alonso M, Sebastian-Leon P, Pellicer A, Valbuena D, Simón C. Window of implantation transcriptomic stratification reveals different endometrial subsignatures associated with live birth and biochemical pregnancy. Fertil Steril 2017;108:703–710.e3.

Diaz-Gimeno P, Sebastian-Leon P, Sanchez-Reyes JM, Spath K, Aleman A, Vidal C, Devesa-Peiro A, Labarta E, Sánchez-Ribas I, Ferrando M, et al. Identifying and optimizing human endometrial gene expression signatures for endometrial dating. Human Reproduction 2022;37:284–296.

Diaz-Gimeno P, Sebastian-Leon P, Spath K, Marti-Garcia D, Sanchez-Reyes JM, Vidal MDC, Devesa-Peiro A, Sanchez-Ribas I, Martinez-Martinez A, Pellicer N, et al. Predicting risk of endometrial failure: a biomarker signature that identifies a novel disruption independent of endometrial timing in patients undergoing hormonal replacement cycles. Fertil Steril 2024;122:352–364.

Dobin A, Davis CA, Schlesinger F, Drenkow J, Zaleski C, Jha S, Batut P, Chaisson M, Gingeras TR. STAR: ultrafast universal RNA-seq aligner. Bioinformatics 2013;29:15–21.

Enciso M, Carrascosa JP, Sarasa J, Martínez-Ortiz PA, Munné S, Horcajadas JA, Aizpurua J. Development of a new comprehensive and reliable endometrial receptivity map (ER Map/ER Grade) based on RT-qPCR gene expression analysis. Human Reproduction 2018;33:220–228.

Evans GE, Phillipson GTM, Sykes PH, McNoe LA, Print CG, Evans JJ. Does the endometrial gene expression of fertile women vary within and between cycles? Hum Reprod 2018;33:452–463.

Fasold M, Binder H. Variation of RNA Quality and Quantity Are Major Sources of Batch Effects in Microarray Expression Data. Microarrays (Basel) 2014;3:322–339.

Garcia-Alonso L, Handfield L-F, Roberts K, Nikolakopoulou K, Fernando RC, Gardner L, Woodhams B, Arutyunyan A, Polanski K, Hoo R, et al. Mapping the temporal and spatial dynamics of the human endometrium in vivo and in vitro. Nat Genet 2021;53:1698–1711.

[dataset] Genome Reference Consortium. 2009. Human Genome Assembly GRCh37. Genome Reference Consortium/NCBI Assembly. GenBank assembly accession: GCA_000001405.1; RefSeq assembly accession: GCF_000001405.13

Hamamah S, Delphine H. Win test.

Hao Y, Stuart T, Kowalski MH, Choudhary S, Hoffman P, Hartman A, Srivastava A, Molla G, Madad S, Fernandez-Granda C, et al. Dictionary learning for integrative, multimodal and scalable single-cell analysis. Nat Biotechnol 2024;42:293–304.

Koot YEM, Van Hooff SR, Boomsma CM, Van Leenen D, Groot Koerkamp MJA, Goddijn M, Eijkemans MJC, Fauser BCJM, Holstege FCP, Macklon NS. An endometrial gene expression signature accurately predicts recurrent implantation failure after IVF. Sci Rep 2016;6:19411.

Liao Y, Smyth GK, Shi W. featureCounts: an efficient general purpose program for assigning sequence reads to genomic features. Bioinformatics 2014;30:923–930.

Parraga-Leo A, Sebastian-Leon P, Devesa-Peiro A, Marti-Garcia D, Pellicer N, Remohi J, Dominguez F, Diaz-Gimeno P. Deciphering a shared transcriptomic regulation and the relative contribution of each regulator type through endometrial gene expression signatures. Reprod Biol Endocrinol 2023;21:84.

Ritchie ME, Phipson B, Wu D, Hu Y, Law CW, Shi W, Smyth GK. limma powers differential expression analyses for RNA-sequencing and microarray studies. Nucleic Acids Res 2015;43:e47.

Sanchez-Reyes JM, Parraga-Leo A, Sebastian-Leon P, Vidal MDC, Marti-Garcia D, Spath K, Sanchez-Ribas I, Sanz FJ, Pellicer N, Remohi J, et al. Stratifying IVF population endometria using a prognosis gradient independent of endometrial timing. Human Reproduction 2025:deaf156.

Seshadri S, Morris G, Serhal P, Saab W. Assisted conception in women of advanced maternal age. Best Pract Res Clin Obstet Gynaecol 2021;70:10–20.

Shi C, Han HJ, Fan LJ, Guan J, Zheng XB, Chen X, Liang R, Zhang XW, Sun KK, Cui QH, et al. Diverse endometrial mRNA signatures during the window of implantation in patients with repeated implantation failure. Hum Fertil (Camb) 2018;21:183–194.

Suhorutshenko M, Kukushkina V, Velthut-Meikas A, Altmäe S, Peters M, Mägi R, Krjutškov K, Koel M, Codoñer FM, Martinez-Blanch JF, et al. Endometrial receptivity revisited: endometrial transcriptome adjusted for tissue cellular heterogeneity. Hum Reprod 2018;33:2074–2086.

Teh WT, Chung J, Donoghue JF, Holdsworth-Carson SJ, Rozen G, Stern C, Rogers PAW. Investigating cycle-to-cycle variation and the impact of endometrial injury through differential gene expression analysis. Hum Reprod 2026:deag025.

Tsoucas D, Dong R, Chen H, Zhu Q, Guo G, Yuan G-C. Accurate estimation of cell-type composition from gene expression data. Nat Commun 2019;10:2975.

Wang W, Vilella F, Alama P, Moreno I, Mignardi M, Isakova A, Pan W, Simon C, Quake SR. Single-cell transcriptomic atlas of the human endometrium during the menstrual cycle. Nat Med 2020;26:1644–1653.

[dataset] Wang W, Vilella F, Alama P, Moreno I, Mignardi M, Isakova A, Pan W, Simon C, Quake SR. 2020. Single cell RNA-seq analysis on human endometrium across the natural menstrual cycle. Gene Expression Omnibus. GSE111976.

de Ziegler D, Cornel C, Bergeron C, Hazout A, Bouchard P, Frydman R. Controlled preparation of the endometrium with exogenous estradiol and progesterone in women having functioning ovaries. Fertil Steril 1991;56:851–855.

