## Supplementary figures and images for "Transcriptomic Reproducibility in Endometrial Biopsies: Evidence Across Cycles and Tissue Regions in Hormonal Replacement Therapy Endometrial Preparation Protocols"

### Supplementary Figure S1

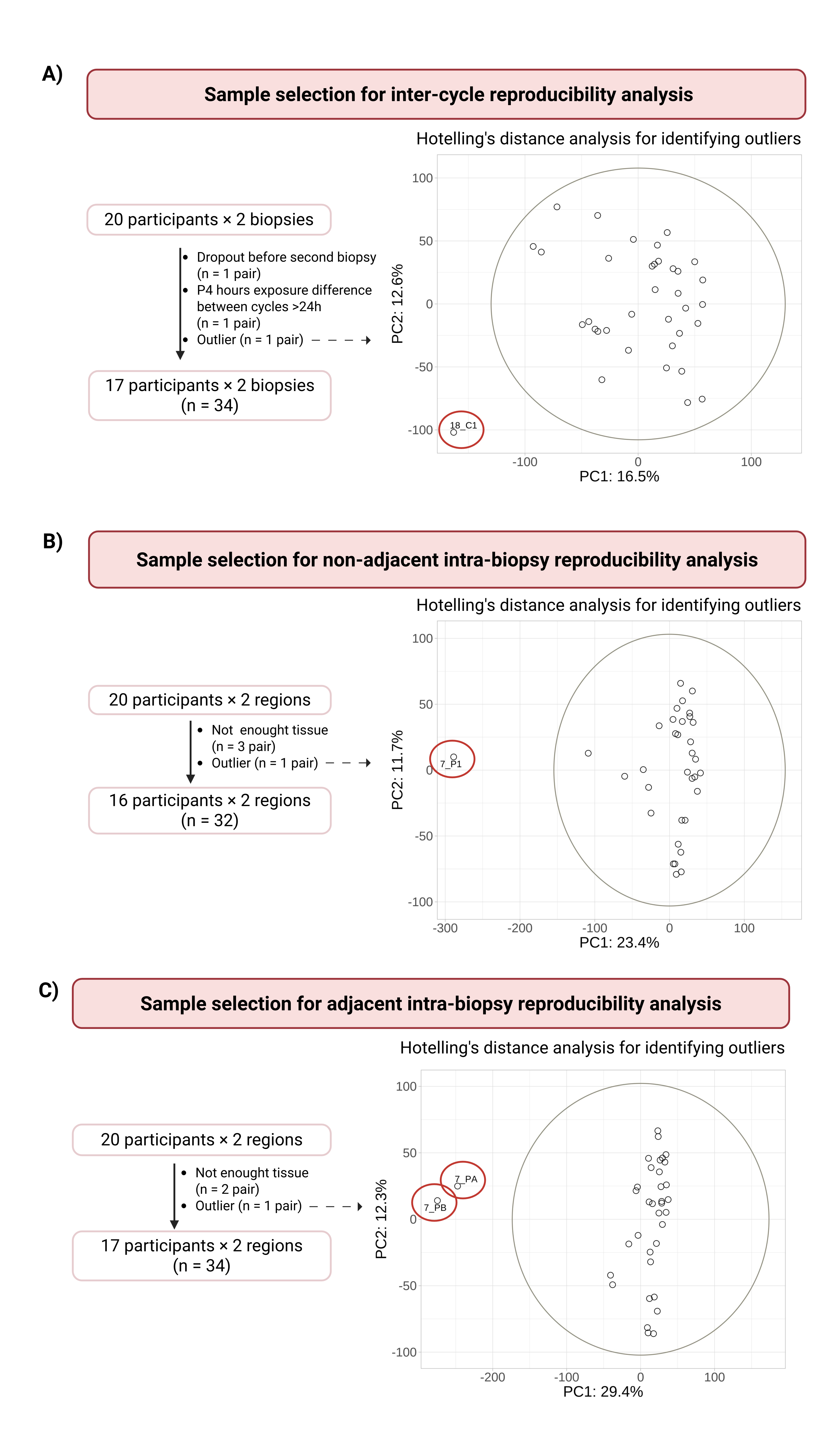

### Supplementary Figure S2

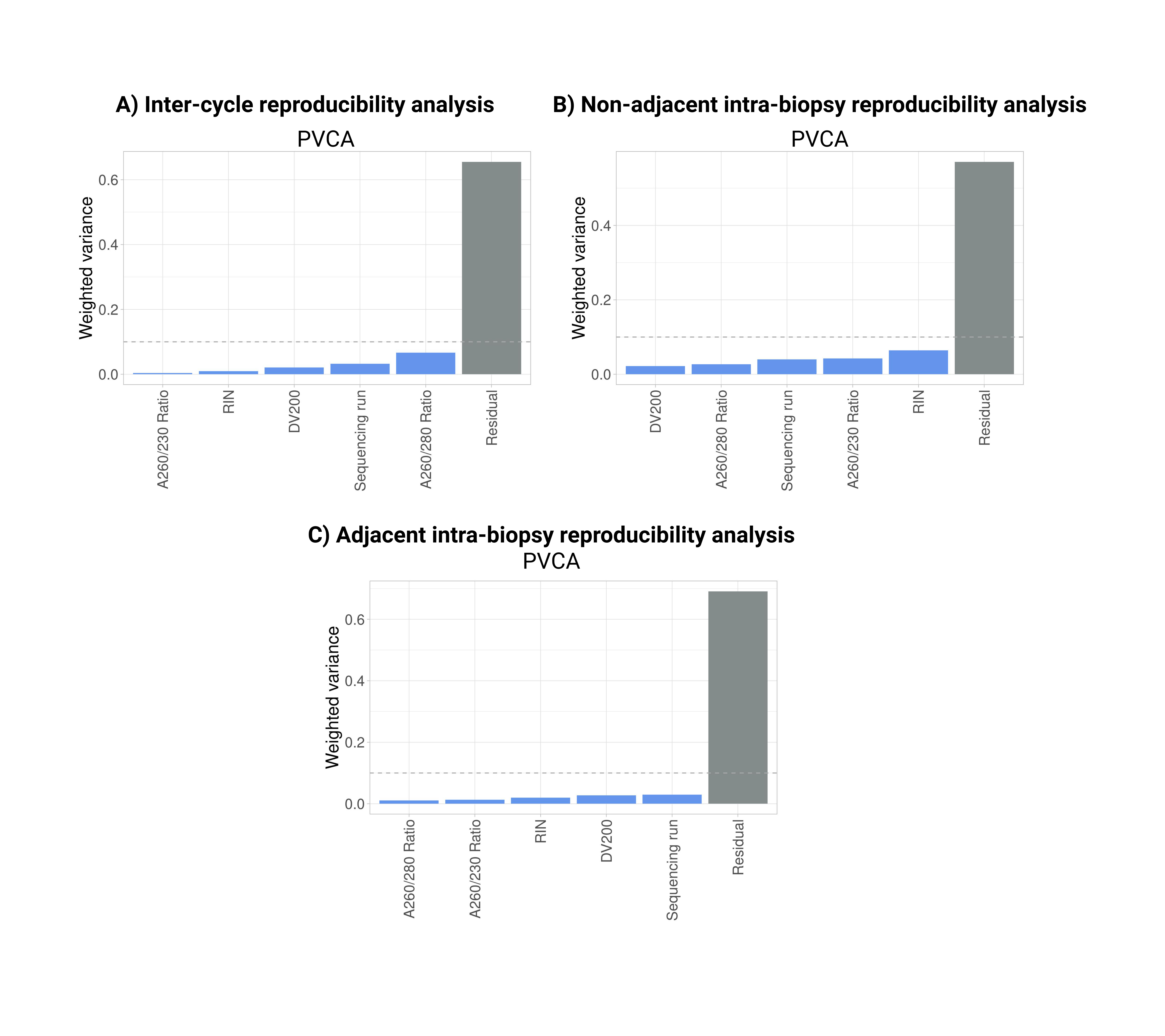

### Supplementary Figure S3

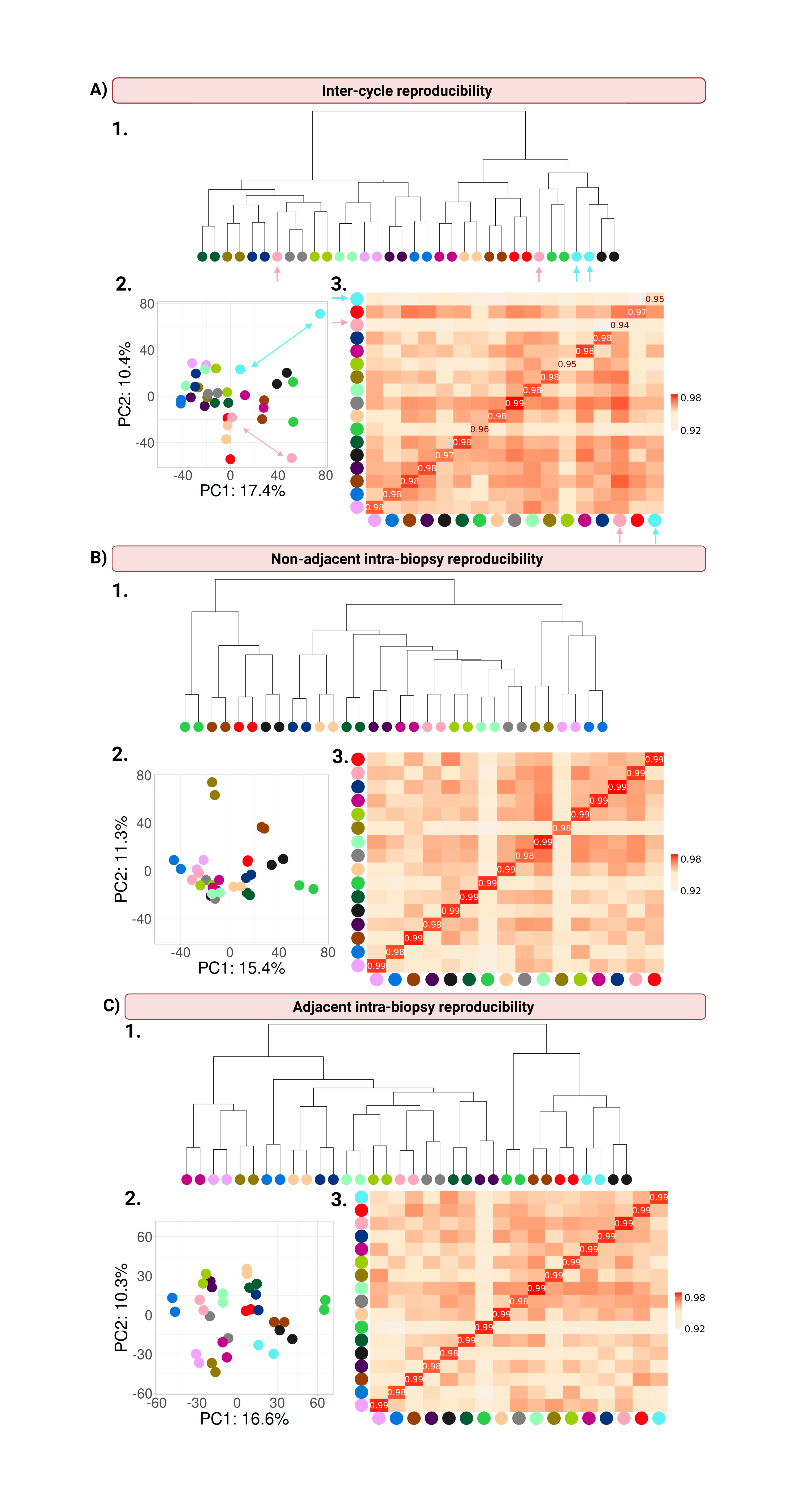
