## Supplementary figures for "Transcriptomic Reproducibility in Endometrial Biopsies: Evidence Across Cycles and Tissue Regions in Hormonal Replacement Therapy Endometrial Preparation Protocols"

**SUPPLEMENTARY MATERIAL**

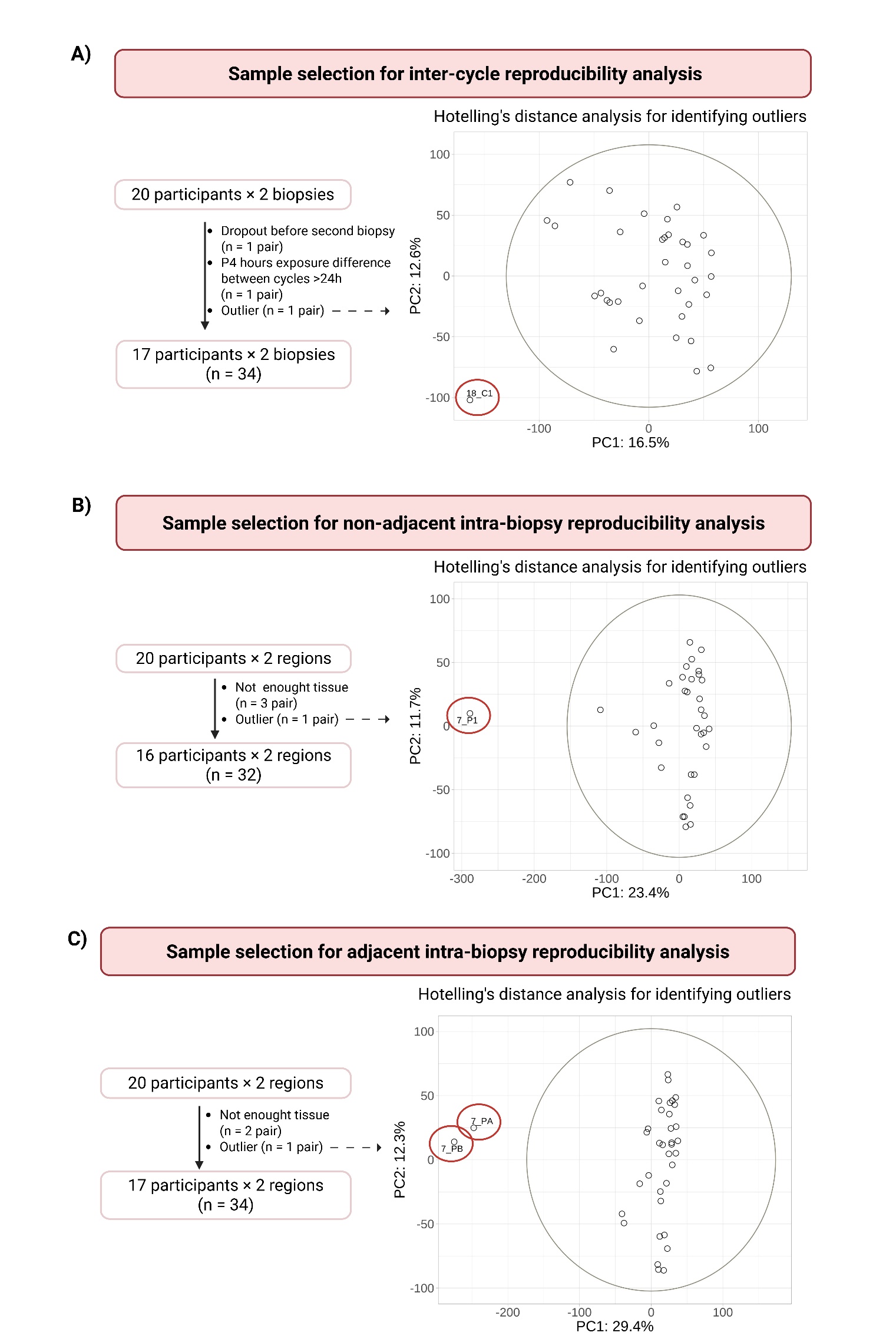

***Supplementary Figure S1. Sample selection.*** *Sample selection and preanalytical processing for the three comparisons performed in this study. For each comparison, 20 participants were initially included. For the inter-cycle comparison* ***(A)****, one participant dropped out before collection of the second biopsy, one biopsy pair showed a difference in progesterone (P4) exposure greater than 24 hours between cycles, and one sample was identified as an outlier based on Hotelling’s distance analysis. After excluding these samples and their corresponding pairs, 17 participants were included in this comparison, comprising a total of 34 samples. For the non-adjacent intra-biopsy comparison* ***(B)*** *and adjacent intra-biopsy comparison* ***(C)****, three and two biopsy pairs, respectively, had insufficient tissue to perform the corresponding analysis. In addition, one sample was identified as an outlier based on Hotelling’s distance analysis in each comparison. After excluding these samples and their corresponding pairs, 16 participants were included in the non-adjacent intra-biopsy comparison, comprising 32 samples, and 17 participants were included in the adjacent intra-biopsy comparison, comprising 34 samples.*

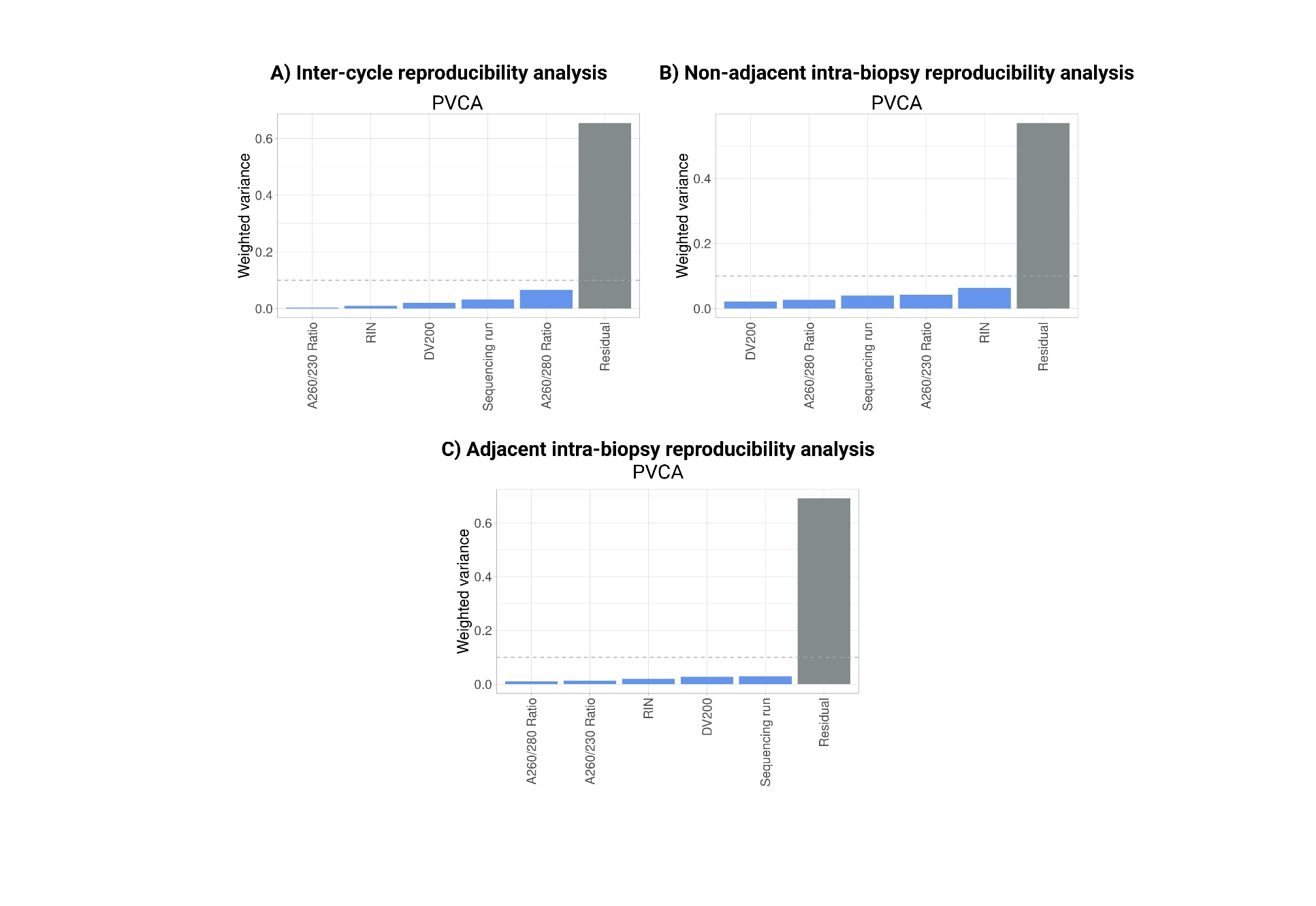

***Supplementary Figure S2. Principal variance component analysis (PVCA) for identifying potential batch effects.*** *Potential batch effects included the A260/230 ratio A260/280 ratio, RNA integrity number (RIN), RNA fragments with more than 200 nucleotides (DV200), and sequencing run. No variable exceeded the predefined 10% variance threshold (gray dotted line) in any of the three reproducibility analyses; therefore, no batch effect correction was required.*

*DV200, RNA fragments with more than 200 nucleotides; RIN, RNA integrity number.*

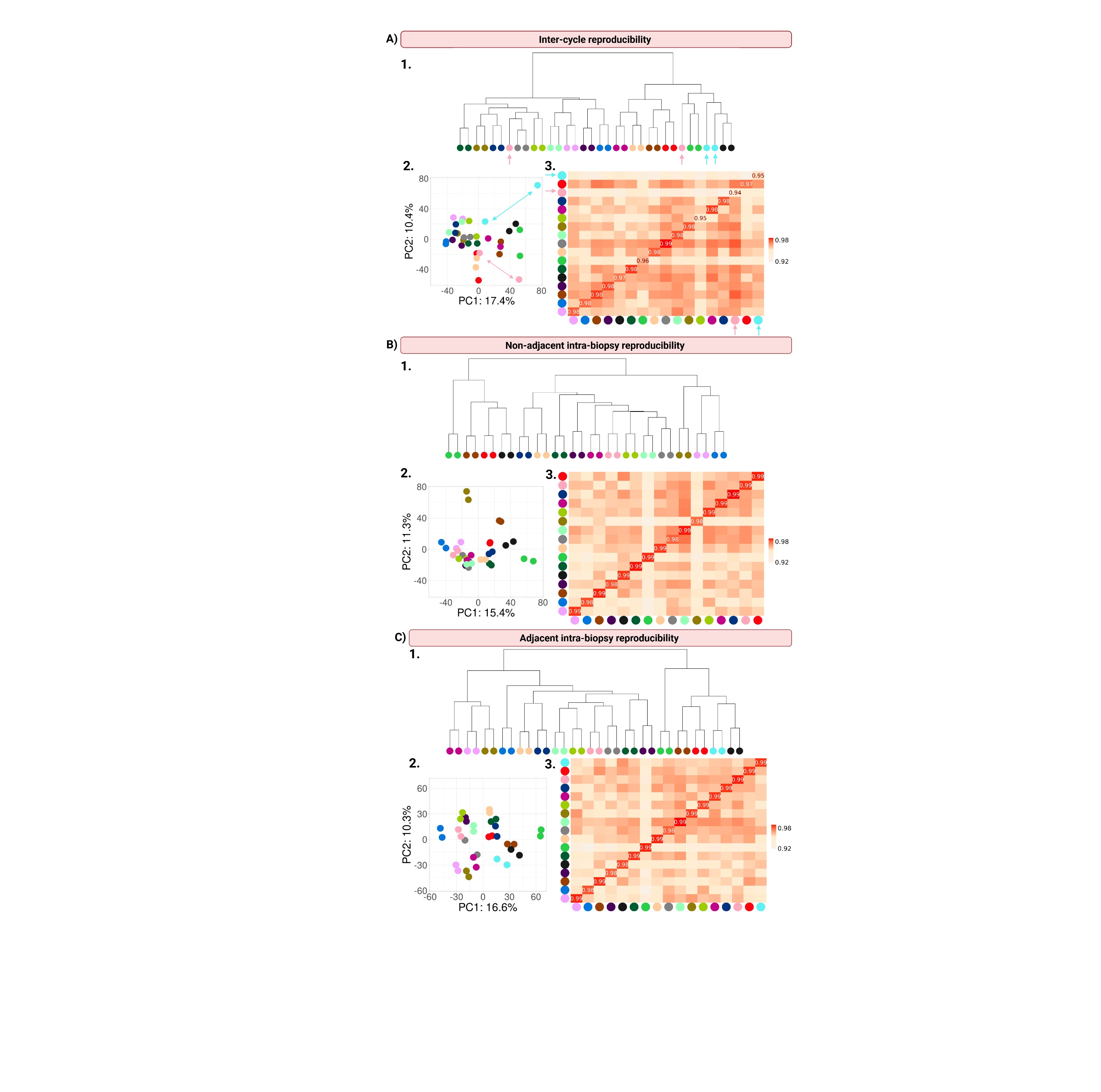

***Supplementary Figure S3. Evaluation of the transcriptomic proximity of paired samples by hierarchical clustering analysis, PCA, and correlation analysis.*** *Within participant/biopsy pairs are marked with the same color. For* *inter-cycle reproducibility* ***(A)*** *most paired samples clustered together in the hierarchical clustering analysis* ***(1)*** *and in the PCA* ***(2)****, with the exception of the blue and pink pairs (indicated with arrows). One sample of each of these pairs also had lower correlation values than the rest of the samples in the correlation analysis with its corresponding pair and with rest of the samples* ***(3)****. For* ***(B)*** *Non-adjacent intra-biopsy reproducibility and* ***(C)*** *adjacent intra-biopsy reproducibility, all samples clustered together and had higher correlation values between pairs.*
