## Supplementary Table S1 for "Transcriptomic Reproducibility in Endometrial Biopsies: Evidence Across Cycles and Tissue Regions in Hormonal Replacement Therapy Endometrial Preparation Protocols"

***Supplementary Table S1.*** *Comparison of cell-type proportions showing no significant differences (false discovery rate [FDR] > 0.05) between paired samples for any cell type.*

| **Comparison** | **Cell type** | ***p* value** | **FDR-adjusted *p* value** |
| --- | --- | --- | --- |
| Inter-cycle | Ciliated | 0.79 | 0.87 |
|  | Endothelia | 0.52 | 0.84 |
|  | Lymphocytes | 0.87 | 0.87 |
|  | Macrophages | 0.29 | 0.78 |
|  | Smooth muscle cells | 0.52 | 0.84 |
|  | Stromal fibroblasts | 0.73 | 0.87 |
|  | Unciliated epithelia 1 | 0.23 | 0.78 |
|  | Unciliated epithelia 2 | 0.14 | 0.78 |
| Non-adjacent intra-biopsy | Ciliated | 0.10 | 0.40 |
|  | Endothelia | 0.35 | 0.56 |
|  | Lymphocytes | 0.54 | 0.72 |
|  | Macrophages | 0.09 | 0.40 |
|  | Smooth muscle cells | 0.85 | 0.85 |
|  | Stromal fibroblasts | 0.31 | 0.56 |
|  | Unciliated epithelia 1 | 0.29 | 0.56 |
|  | Unciliated epithelia 2 | 0.79 | 0.85 |
| Adjacent intra-biopsy | Ciliated | 0.70 | 0.80 |
|  | Endothelia | 0.06 | 0.24 |
|  | Lymphocytes | 0.21 | 0.57 |
|  | Macrophages | 0.02 | 0.19 |
|  | Smooth muscle cells | 0.69 | 0.80 |
|  | Stromal fibroblasts | 0.42 | 0.68 |
|  | Unciliated epithelia 1 | 0.35 | 0.68 |
|  | Unciliated epithelia 2 | 0.93 | 0.93 |
